# Iron-Oxide Nanoparticle MRI for Human Brain Tumors: A Systematic Review and Protocol-Level Meta-analysis of Administered Doses Across Ferumoxtran-10, Ferumoxytol, and Ferumoxides

**DOI:** 10.64898/2025.12.07.25341771

**Authors:** Farzan Fahim, Negin Safari Dehnavi, Mahsa Hemmati, Mohammadamin Sabbagh Alvani, Ali Khorram, Ali Saravani, Ali Rezaeian, Muhammad Parsa Pashazadeh, Ibrahim Mohammadzadeh, Sayeh Oveisi, Saeed Oraee-Yazdani, Alireza Zali

## Abstract

**Background:** Gadolinium-based contrast agents (GBCAs) have recognized limitations for accurate delineation of brain tumor margins and perfusion assessment in neuro-oncology. Nanoparticle contrast agents (NP-CAs), particularly ultrasmall superparamagnetic iron oxides (USPIOs), may overcome these limitations by providing delayed uptake and tissue characterization.

**Methods:** We systematically searched PubMed, Embase, Scopus, Web of Science, and trial registries on July 2025 for human neuro-oncology studies using NP-CAs. The primary outcome was the change in relative cerebral blood volume (ΔrCBV) compared with that of GBCA imaging at prespecified time points. The secondary outcomes were the contrast-to-noise ratio (CNR), signal-to-noise ratio (SNR), reader-rated margin delineation, and safety. Two reviewers independently extracted the data and assessed the risk of bias via tools from the Joanna Briggs Institute. Random effects meta-analysis was performed when ≥3 comparable datasets were available; otherwise, the results were synthesized narratively.

**Results:** Six studies met the inclusion criteria. Agents included ferumoxtran-10, ferumoxytol, and ferumoxides, with intravenous doses ranging from 0.56–7.00 mg Fe/kg. The pooled common-effect mean dose was 4.65 mg/kg. Across heterogeneous designs, NP-CAs consistently enhanced margin delineation and perfusion metrics: ferumoxtran-10 produced sharp, persistent T2/T2* rims beyond T1-GBCA enhancement, and ferumoxytol-based DSC (Dynamic Susceptibility Contrast) yielded a higher rCBV with reduced leakage effects. No serious adverse events were reported; infusion reactions were rare and inconsistently defined.

**Conclusions:** Compared with GBCA, NP-CAs, particularly USPIOs, improve brain tumor margin visualization and perfusion assessment. However, methodological heterogeneity and small sample sizes limit certainty. Standardized protocols for dosing, acquisition, and safety monitoring, alongside biopsy-validated prospective trials, are needed before clinical adoption.

## Introduction

Brain tumors, including gliomas, meningiomas, and pituitary tumors, account for more than 120 histological subtypes and remain a major cause of neurological morbidity and mortality despite advances in neurosurgery, oncology, and radiotherapy [1,2]. Patients often present with cognitive deficits, focal neurological impairments, or behavioral changes that compromise functional independence [3–5]. Early and accurate diagnosis is critical, as timely intervention can improve both survival and quality of life [2,5].

Magnetic resonance imaging (MRI) is the standard tool for the diagnosis, treatment planning, and longitudinal monitoring of brain tumors [6,7]. Contrast-enhanced T1-weighted MRI using gadolinium-based contrast agents (GBCA) is widely employed, particularly for glioma evaluation [8–11]. However, GBCA has inherent limitations, such as variable blood–brain barrier (BBB) permeability, the absence of tumor-specific binding, and a tendency to underestimate infiltrative tumor margins, which can result in subtotal resections and suboptimal targeting in radiotherapy [11–13].

Nanoparticle-based MRI contrast agents (NP-CAs), especially ultrasmall superparamagnetic iron oxides (USPIOs), offer alternatives with distinct biological and imaging properties [12,14–19,22]. Owing to their small size (typically 20–60 nm) and modifiable surface chemistry, they can cross the BBB, accumulate via passive enhanced permeability and retention (EPR) or active ligand targeting, and induce strong susceptibility effects on T1/T2* sequences [16,18–20,38]. Unlike GBCAs, USPIOs permit both perfusion imaging during early intravascular phases and margin visualization on delayed imaging for up to 24–36 hours, highlighting immune-rich or infiltrated tumor boundaries [27–33]. Preclinical studies also suggest potential theranostic applications, including targeted drug delivery and immune modulation [21,22].

While multiple small clinical reports have described these advantages, evidence on diagnostic performance, reproducibility, safety, and comparative effectiveness versus GBCA remains fragmented [22,27–33,35,36]. Moreover, no prior synthesis has quantified dosing patterns or examined protocol heterogeneity in human neuro-oncology.

We therefore performed a systematic review of human studies assessing NP-CAs in brain tumor MR images, with a primary focus on changes in relative cerebral blood volume (ΔrCBV) compared with the GBCA at prespecified time points. The secondary outcomes included the contrast-to-noise ratio (CNR), signal-to-noise ratio (SNR), reader-rated tumor margin delineation, and safety.

## Methods

### Registration, protocol, and reporting

This review followed the PRISMA 2020 guidelines [23, 24]. The protocol was registered in PROSPERO (CRD420251106029) before the start of screening. Two pragmatic protocol deviations were introduced prior to data extraction:

1. Single-patient case reports were retained in the qualitative synthesis owing to limited human evidence.
2. As harmonized segmentation metrics (e.g., Dice and Hausdorff) are rarely available, we prioritized extractable quantitative MRI readouts (signal intensity change, SNR, CNR, and rCBV) alongside reader-reported lesion and border delineation.
3. Title was updated to “Iron-Oxide Nanoparticle MRI for Human Brain Tumors: A Systematic Review and Protocol-Level Meta-analysis of Administered Doses Across Ferumoxtran-10, Ferumoxytol, and Ferumoxides” to emphasize on Iron-Oxide nanoparticles and the meta-analysis of administered doses across ferumoxtran-10, frumoxytol, and ferumoxide. This change was made to reflect the focus of the included studies.

The PRISMA checklist, full protocol, database search strings, and data extraction form are provided in the Supplementary Materials1 to 4 respectively.

### Information sources and search strategy

We searched PubMed, Embase, Web of Science Core Collection, Scopus, and the Cochrane trial Library from database inception to 12 July 2025. No date or language restrictions were applied. We also perform backward and forward citation chasing. The full line-by-line strategies appear verbatim in the Supplementary materials 3.

### Keyword frameworks

#### PubMed (MeSH + text)

(“Nanoparticles”[MeSH] OR nanoparticle* OR “superparamagnetic iron oxide” OR SPIO OR USPIO OR ferumoxytol OR “gold nanoparticle*” OR “quantum dot*”) AND (“Contrast Media”[MeSH] OR “contrast agent*”) AND (“Magnetic Resonance Imaging”[MeSH] OR MRI) AND (brain OR intracranial OR glioma* OR glioblastoma OR metastas*).

#### Embase (Emtree + text)

(‘nanoparticle’/exp OR nanoparticle*:ti, ab OR ‘superparamagnetic iron oxide’/exp OR ferumoxytol) AND (‘contrast medium’/exp OR ‘contrast agent*’:ti, ab) AND (‘magnetic resonance’/exp OR MRI:ti, ab) AND (brain:ti, ab OR intracranial:ti, ab OR glioma*:ti, ab).

#### Web of Science (Topic)

TS=(nanoparticle* OR “superparamagnetic iron oxide” OR ferumoxytol) AND TS=(contrast NEAR/2 (agent* OR media)) AND TS=(MRI OR “magnetic resonance”) AND TS=(brain OR glioma* OR metastas*).

#### Scopus (TITLE-ABS-KEY)

TITLE-ABS-KEY (nanoparticle* OR “superparamagnetic iron oxide” OR ferumoxytol) AND TITLE-ABS-KEY (contrast AND (agent* OR media)) AND TITLE-ABS-KEY (MRI OR “magnetic resonance”) AND TITLE-ABS-KEY (brain OR glioma* OR metastas*).

#### Cochrane Library

(nanoparticle* OR ferumoxytol OR “superparamagnetic iron oxide”) AND (MRI OR “magnetic resonance”) AND (brain OR glioma*).

### Study selection

Titles/abstracts and then full texts were screened independently by AS and AK; disagreements were resolved by a third reviewer (FF). The PRISMA flow diagram for selection: 324 records identified; 149 removed before screening; 175 titles/abstracts screened with 169 exclusions, mostly animal studies (n= 119), reviews (n= 43), in vitro work (n= 6), and not related (n= 1). We retrieved all the full texts; six studies met the inclusion criteria and were included. The database-level reasons for exclusion are shown in Figure 1.

**Figure 1.**
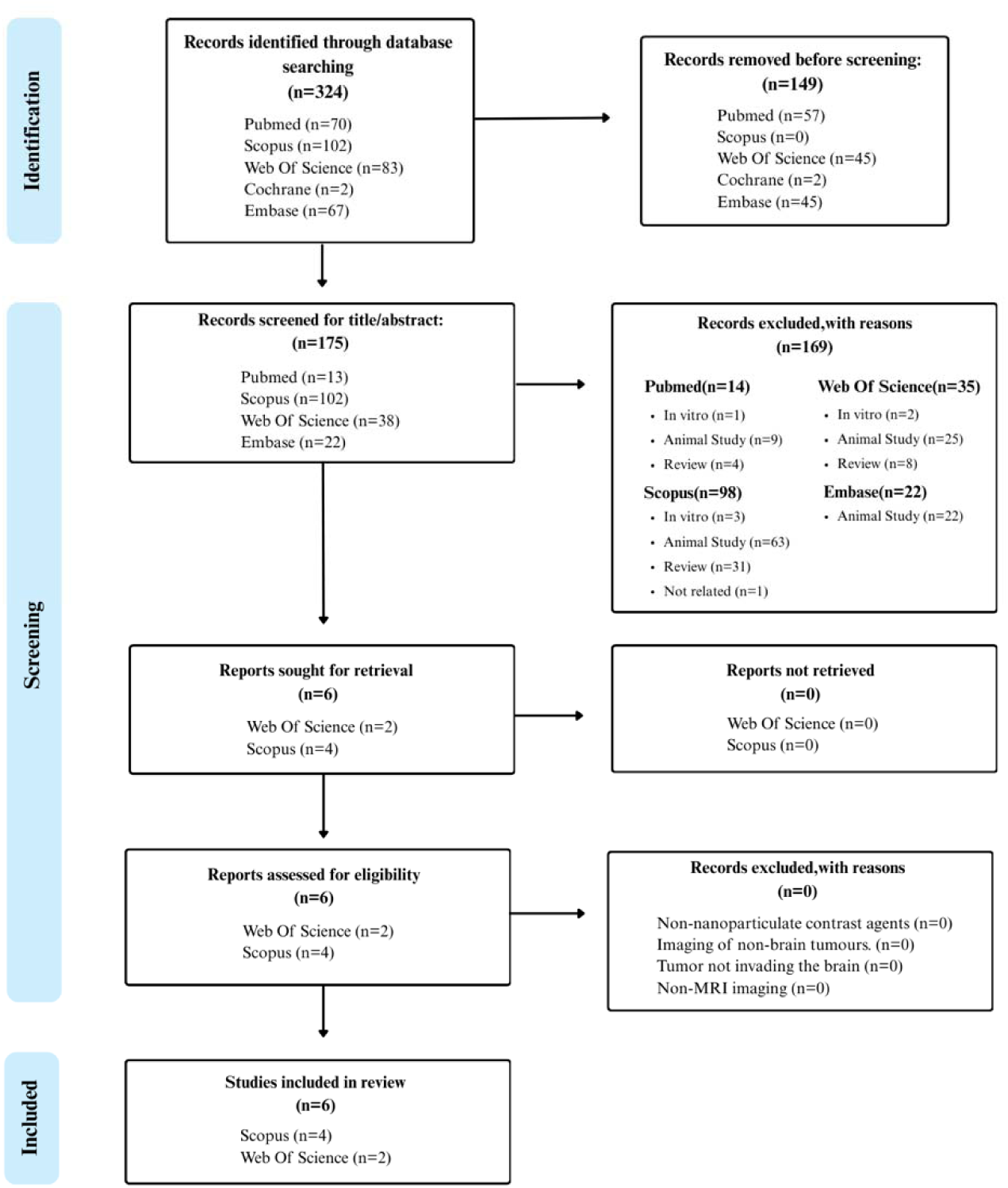
**PRISMA 2020 flow diagram** – study selection process

### Eligibility criteria

#### Population

Human participants with primary or secondary brain tumors who underwent MRI after the administration of a nanoparticle-based contrast agent.

#### Intervention and comparator

MRI enhanced with any nanoparticle contrast agent compared with conventional gadolinium-based agents or non-contrast MRI. When available, histopathology or intraoperative findings were used as reference standards.

### Outcomes

#### Primary

Mean difference in the ΔrCBV between NP-CA imaging and gadolinium-enhanced MRI at prespecified time points.

#### Secondary

CNR, SNR, reader-rated tumor margin delineation, and safety/adverse events.

##### Study designs

Randomized trials, prospective or retrospective cohorts, and case series (≥ 2 patients) were eligible for quantitative synthesis if variance data were available (SD, SE, or per-patient values enabling calculation).

##### Exclusions

In vitro or animal studies, non-MRI imaging modalities, abstracts-only publications, reviews, editorials, and duplicate reports.

### Data extraction

Using a piloted, standardized form, **NSD** and **AS** independently extracted: study identifiers (authors, year, country, design), tumor type, grade, sample size, MRI platform and sequences, nanoparticle characteristics (material, coating/functionalization, dose, route, timing), comparators, reference standards, quantitative outcomes (SI change, SNR, CNR, rCBV), reader-reported delineation, safety, and analysis methods; funding and sponsorship were recorded and reported. Discrepancies were resolved by consensus. The study-level extraction variables and raw metrics are provided in supplementary material 4.

### Risk of bias assessment

We assessed methodological quality with the Joanna Briggs Institute (JBI) Critical Appraisal Checklists tailored to study design (cohort, case series, and case reports)[25]. NSD and AK completed the assessments independently; AR adjudicated disagreements. For each study, we summarized the JBI percentage score and classified overall risk as high (20–49%), moderate (50–79%), or low (80–100%). The checklists consider, among other items, clear inclusion criteria; appropriate sampling/recruitment; similarity of groups (where applicable); valid and reliable exposure/outcome measurement; identification and management of confounders; completeness of follow-up; and appropriate statistical analysis[26]. The study-level scores and predominant bias domains are summarized in Figure 2. Item-level checklists are provided in the supplementary materials 5.

**Figure 2.**
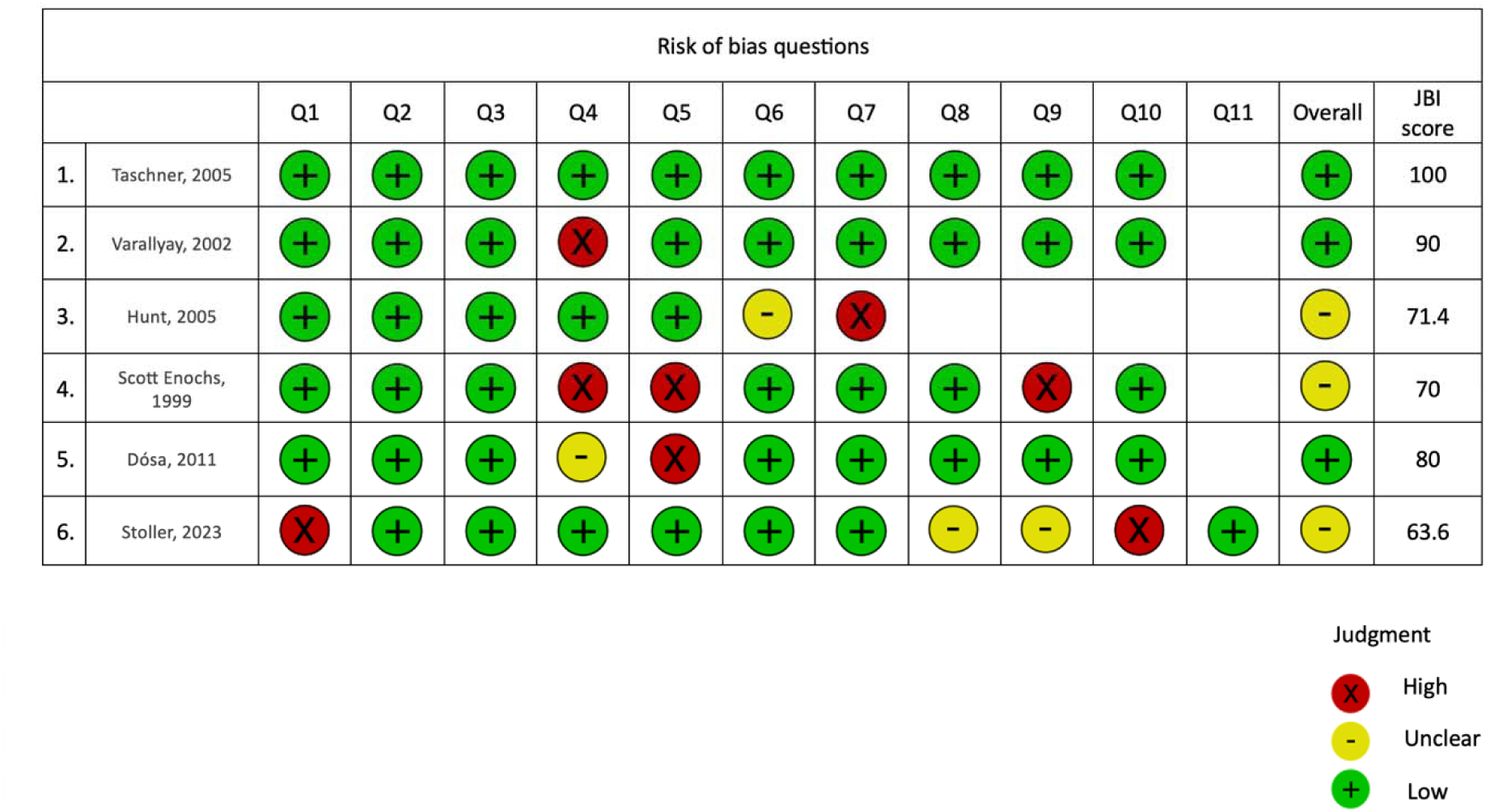
**Risk of bias assessment-** performed via the Joanna Briggs Institute (JBI) critical appraisal tool.

### Data synthesis and statistical analysis

We conducted a single-arm meta-analysis to pool the per-protocol nanoparticle iron doses (mg Fe/kg) reported in studies where all participants received the same fixed dose. From each study, we extracted the study label, sample size (N), and administered dose. Because within-study variability is essentially protocol noise (all participants receive one fixed dose), we assumed a per-participant SD of 0.10 mg/kg (with a planned sensitivity range of 0.05–0.20 mg/kg). Standard errors were computed as SD/√N, and studies were synthesized with metagen() from the R package meta, using inverse-variance weighting. We report both common-effect and random-effects models; between-study variance (τ²) was estimated with REML, and random-effects CIs were estimated via Hartung–Knapp adjustment. Forest and funnel plots were generated, with studies sorted by their mean dose; no prediction interval was reported.

No quantitative meta-analysis of prespecified imaging outcomes (ΔrCBV, CNR, SNR, or reader-rated margin delineation) was feasible. Across the six included studies (n=68), only one study (Dósa 2011) reported extractable rCBV values; CNR/SNR values were either absent or reported without variance; and margin delineation was uniformly qualitative (without numerical scales). Because no outcome provided ≥3 datasets with compatible units and variance measures (SD, SE, or per-patient values), quantitative pooling was not possible in accordance with the PRISMA 2020 and PROSPERO protocol requirements. Therefore, the only analyzable variable was the fixed-dose regimen, which is a protocol-level parameter with inherently minimal within-study variability.

## Results

### Study Selection and Characteristics

Of the 324 records identified, six human studies published between 1999 and 2023 (five from the USA and one from Switzerland) met the eligibility criteria. The study designs included three multi-patient case series, two prospective cohorts, and one case report. The sample sizes ranged from 2 to 26, with a total of n = 68 participants. The tumor types included both primary and secondary lesions, namely, WHO grade II–IV gliomas, gliosarcoma, lymphoma, metastases (Non-Small Cell Lung Cancer (NSCLC), melanoma), meningioma, and pituitary adenoma. One recent study (Stoller 2023) incorporated stereotactic biopsies for histopathological correlation. Summary of included studies is presented in table-1.

**Table 1.** Summary of included studies.

| Study (Year, Country) | Design | n (mean age; % female) | Tumor types | Agent (type) | Dose & regimen | MRI timing & sequences | Main findings vs Gd | Safety | JBIRisk |
| --- | --- | --- | --- | --- | --- | --- | --- | --- | --- |
| Enochs 1999 (US) | Case series | 4 (40 y; 0%) | Gliomas; melanoma metastasis | Ferumoxtran-10 (USPIO) | 1.1 mg/kg, single dose | 1.5 T; T1, T2, GRE; 12–36 h post-infusion | Thinner, sharper T2/T2* rims; persisted to 36 h; some Gd-enhancing lesions were USPIO-negative; histology confirmed iron uptake. | NR | Low |
| Varallyay 2002 (US) | Case series | 20 (47.2 y; 40%) | Mixed intracranial tumors | Ferumoxtran-10 (USPIO) vs Ferumoxides (SPIO) | 2.6 mg/kg, single dose | 1.5 T; SE T1, fast SE T2, GRE T2; 6–24 h post-infusion | Ferumoxtran-10 produced sharp, persistent rims exceeding Gd; Ferumoxides showed minimal uptake. | NR | Low |
| Taschner 2005 (Switzerland) | Case series | 9 (62 y; 33%) | Gliosarcoma; ependymoma; metastasis; lymphoma | Ferumoxtran-10 (USPIO) | 2.6 mg/kg, single dose | 1.5 T; T1 SE, T2 TSE, GRE; ~24 h post-infusion | USPIO T2 margins matched or exceeded T1 Gd; absent in necrosis; complements but does not replace Gd. | Well tolerated | Low |
| Hunt 2005 (US) | Case series | 2 (NR; 0%) | Anaplastic oligodendroglioma; GBM | Ferumoxtran-10 (USPIO) | 2.6 mg/kg, single dose | 0.15 T intraoperative; T1, T2, GRE; ~24 h post-infusion | Enabled intraoperative MRI without Gd artifacts; clear tumor margins at low field. | NR | Low |
| Dósa 2011 (US) | Cohort | 26 (47 y; 30.8%) | Glial tumors; lymphoma; metastases; meningioma; adenoma | Ferumoxytol (USPIO) | 510 mg fixed dose | 3 T; T1 SE, T2 TSE, DSC perfusion; minutes–24 h post-infusion | Ferumoxytol yielded higher rCBV and detected inflammation; Gd provided crisper T1 borders. | No SAE | Low |
| Stoller 2023 (US) | Cohort | 7 (61.8 y; 14.3%) | Recurrent GBM | Ferumoxytol (USPIO) | 7.0 mg/kg, single dose | 3 T; 3D T1 TFE, FLAIR; ~24 h post-infusion | Immune phenotyping via susceptibility maps distinct from Gd; biopsy co-localized iron with macrophage-rich regions. | No SAE | Unclear |
| USPIO = ultrasmall superparamagnetic iron oxide; SPIO = superparamagnetic iron oxide; Gd = gadolinium; GRE = gradient-echo; SE = spin-echo; TSE = turbo spin-echo; DSC = dynamic susceptibility contrast; rCBV = relative cerebral blood volume; SAE = serious adverse event; GBM = glioblastoma. |  |  |  |  |  |  |  |  |  |

### Participant Demographics

Across all the studies, the ages ranged from 19–77 years (pooled mean 51.6 y). The sex distribution was heterogeneous, with reported female proportions ranging from 0% to 40%. Summing those with available data resulted in ≥ 20 women (≥ 29.4%). One study did not report sex.

### Nanoparticle Agents and Dosing

The agents used were iron-oxide nanoparticles:

- Ferumoxtran-10 (USPIO): AMI-227/Sinerem/Combidex, dextran-coated, hydrodynamic diameter approximately 20–50 nm.
- Ferumoxytol (SPIO): Feraheme, ∼30 nm.
- Ferumoxides (SPIO): Feridex IV, ∼20–60 nm.

Administrations were intravenous. Weight-based dosing spanned 0.56–7.00 mg Fe/kg; one protocol used a fixed 510 mg ferumoxytol dose. The doses clustered near 2.6 mg/kg (USPIO) and 7.0 mg/kg (high-dose ferumoxytol). Quantitative synthesis of the primary outcome (ΔrCBV) could not be performed because only Dósa 2011 (n=26) provided variance-reported perfusion metrics; Stoller 2023 reported directional changes but without extractable numerical values. CNR/SNR datasets were similarly incomplete (no variance across all studies). Margin delineation was consistently descriptive and unsuitable for statistical pooling. Consequently, per-protocol dose was the only variable meeting minimal criteria for meta-analysis.

### MRI platforms, sequences, and timing

Five studies utilized 1.5 T or 3 T scanners, while one study employed an intraoperative MRI with a field strength of 0.15 T. The sequences included T1-SE/TSE, T2-TSE, GRE, and DSC perfusion. The acquisition timing ranged from first-pass minutes to delayed imaging at 12–36 h post-injection, most often approximately 24 h.

All studies reported directionally favorable effects of NP-CAs compared with gadolinium:

### Imaging Performance (Quantitative and Qualitative)

- Ferumoxtran-10 produces thin, sharp T2/T2* rims that persist for up to 36 h and often extend ≥ 3 mm beyond T1-Gd enhancement.
- Ferumoxytol (DSC) yielded a higher rCBV (+18–25% vs Gd; p<0.05), with less leakage-artifact sensitivity.
- The absence of NP uptake corresponded to regions of necrosis or non-viable tissue (supporting differentiation from progression).

Reported p-values for signal-intensity changes: < 0.001 to 0.015.

### Risk of Bias

Methodological quality was assessed via the Joanna Briggs Institute (JBI) Critical Appraisal Checklist, with the checklist type matched to each study design (case series, cohort, or case report). Two reviewers (NSD, AK) conducted independent assessments, with a third (AR) adjudicating disagreements. Each study received a percentage score based on the proportion of “Yes” responses across applicable items; overall risk categories were assigned as low (80–100%), moderate (50–79%), or high (20–49%) risk of bias.

Across the six eligible human studies (n = 68), five were rated as low risk, and one was rated as unclear risk. Importantly, even the low-risk studies suffered from recurring minor issues, and the unclear-risk study contained multiple unresolved methodological concerns.

### Study-level Appraisals

Enochs 1999 – Low risk

- Strengths: Clear inclusion criteria; detailed description of the patient population (gliomas, metastatic melanoma); standardized NP-CA administration (USPIO, known core/hydrodynamic sizes, dose); precisely reported MRI timing (12, 24, 36 h) and sequences; histopathological confirmation of imaging findings.
- Limitations: Small sample size (n = 4); non-consecutive enrollment; incomplete demographic reporting (sex distribution omitted); limited data on adverse events (AEs).

Varallyay 2002 – Low risk

- Strengths: Well-defined target population; direct within-study comparison of ferumoxtran-10 vs. ferumoxides; explicit dosing; reproducible post-contrast timing; clear outcome definitions (margin sharpness, uptake pattern).
- Limitations: Potential selection bias due to unclear consecutiveness of recruitment and incomplete demographic breakdown.

Taschner 2005 – Low risk (100% of JBI items)

- Strengths: Complete transparency in inclusion/exclusion; precise dosing and infusion duration; uniform imaging protocols; all domains met the JBI criteria; explicit handling of radiation necrosis cases; and full AE reporting.
- Limitations: The sample size was modest (n = 9), reducing the statistical precision, although this did not affect the bias rating.

Hunt 2006 – Low risk

- Strengths: Defined patient/tumor characteristics; protocol adherence for intraoperative low-field MRI; same-day contrast-enhanced imaging with clear depiction of the NP-CA benefits.
- Limitations: Very small sample size (n = 2); absence of any AE reporting.

Dósa 2011 – Low risk

- Strengths: Largest series (n = 26); fixed--dose ferumoxytol protocol; detailed imaging methodology; inclusion of quantitative perfusion metrics (rCBVs); safety assessment.
- Limitations: Non-consecutive patient inclusion not explicitly excluded; incomplete demographic reporting in some sub-groups.

Stoller 2023 – Unclear risk (6/11 JBI items as “Yes”)

- Strengths: Prospective cohort design; within-patient histopathological correlation; standardized ferumoxytol protocol (7 mg/kg); advanced imaging sequences at 3 T.
- Limitations (multiple domains):
  - The recruitment process was not described as consecutive, introducing possible selection bias.
  - Baseline comparability of patients not clearly demonstrated (e.g., prior treatments, disease status).
  - Handling of incomplete follow-up/attrition not reported.
  - Adverse event reporting minimal (limited to statements of no SAEs; no grading of non-serious events).
  - Clinical outcomes post-intervention not characterized.

These unresolved items prevented a low-risk rating despite otherwise modern methodological features.

### Cross-study Patterns of Potential Bias

1. Non-consecutive or unclear recruitment: observed in Enochs, Varallyay, Dósa, Stoller; increases risk of selection bias.
2. Incomplete demographic reporting affects generalizability and subgroup analysis potential.
3. Adverse events under-reporting: especially for non-serious infusion reactions; only some studies quantified and graded events.
4. Small sample sizes: even in low-risk studies, n < 10 in four studies, limiting precision and increasing the likelihood of chance findings.
5. Protocol heterogeneity: differences in NP chemistry, dosing (1.1–7.0 mg/kg), MRI timing (minutes to approximately 36 h), and field strength (0.15 T–3 T) may confound the consistency of effects and reduce certainty.

### Summary Judgment

While the majority of included studies achieved a low-risk rating and demonstrated internally valid methods, selection bias, limited AE documentation, and small, heterogeneous samples remain the most important threats to validity. This single, unclear-risk study highlights the need for transparent recruitment protocols, complete AE reporting, and thorough follow-up in future neuro-oncology NP-CA research.

### Feasibility of Quantitative Outcome Synthesis

Quantitative pooling of the prespecified imaging outcomes was not feasible. Among the six included studies (n = 68), variance-reported ΔrCBV values were available only from Dósa 2011; Stoller 2023 provided directional perfusion changes without extractable numerical values. CNR and SNR were either not reported or were presented without standard deviations or standard errors across all studies. Reader-rated tumor margin delineation was uniformly qualitative and lacked any numerical scoring scale. Because none of these outcomes provided ≥3 datasets with compatible definitions, units, and variance reporting, no imaging endpoint met the PRISMA 2020 criteria for meta-analysis. Therefore, all imaging outcomes are synthesized narratively, and the only variable eligible for quantitative pooling was the per-protocol nanoparticle iron dose.

### Meta-analytic dose findings

Across six studies (N=68), fixed doses ranged from 1.10–7.00 mg Fe/kg, clustering near 2.6 and 7.0 mg/kg. The common-effect pooled mean was 4.65 mg/kg (95% CI 4.62–4.67); the random-effects estimate (REML + Hartung–Knapp) was 3.82 mg/kg (95% CI 1.16–6.48). Between-study heterogeneity was extreme (I²=100%, τ²=6.4166), which is consistent with bimodal protocols. Forest plots (Figure 3) show weighted study estimates; the funnel plot (Figure 4) demonstrates dispersion reflecting the two clusters; with only six studies and protocol-driven clustering, asymmetry is difficult to interpret for small-study effects.

**Figure 3.**
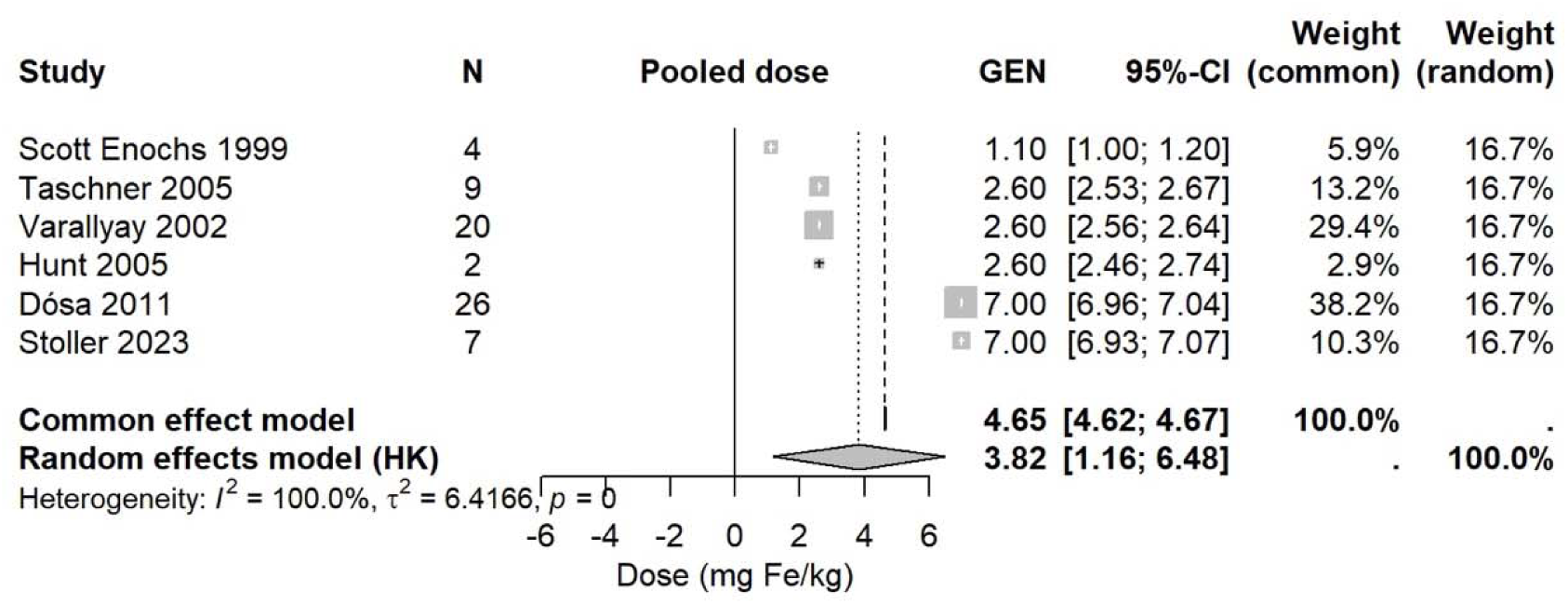
**Forest plot-**Fixed-dose nanoparticle iron

**Figure 4.**
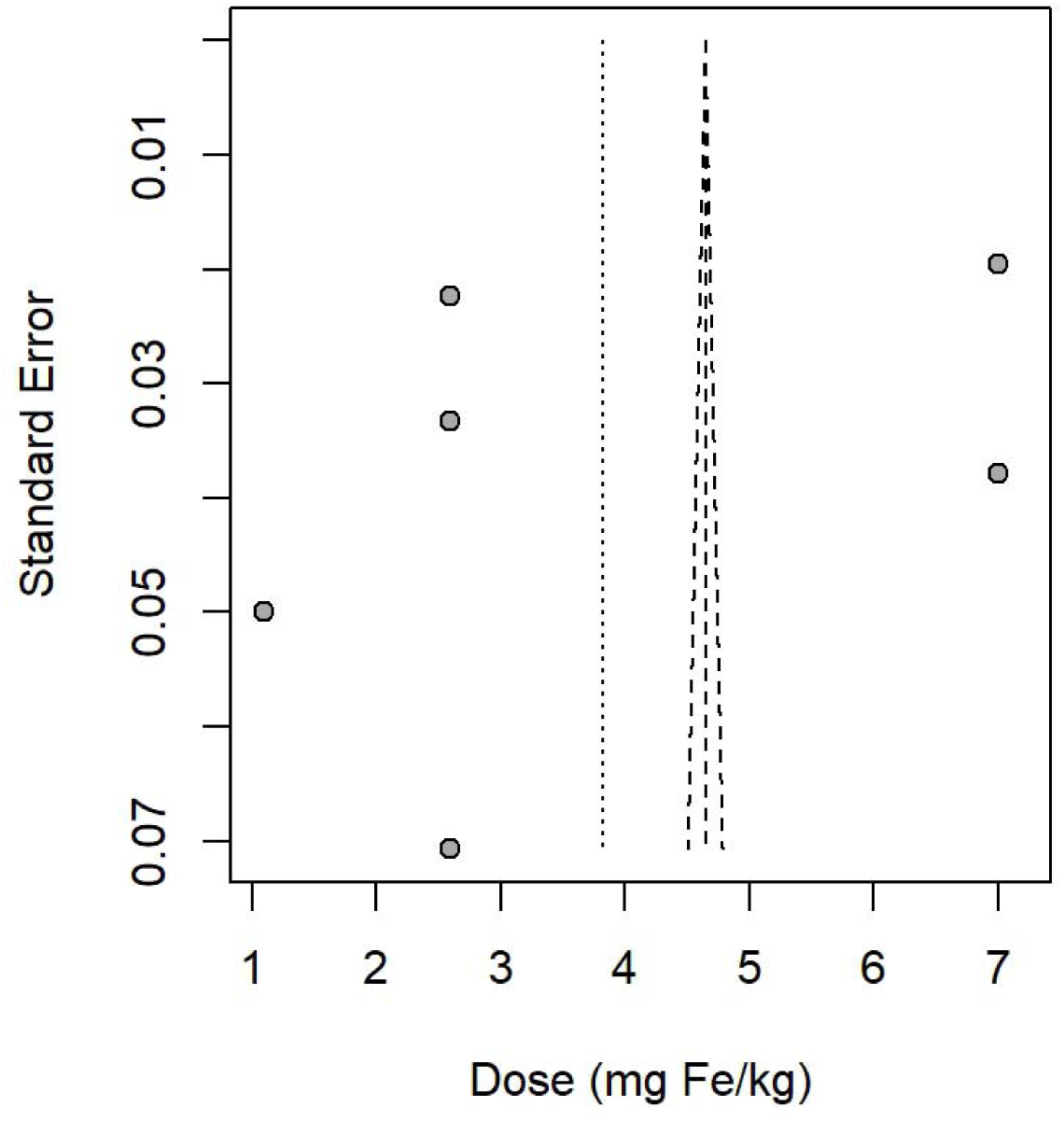
**Funnel plot-**Fixed-dose nanoparticle iron

### Certainty of evidence (GRADE)

The certainty ratings for each prespecified outcome are summarized in Table 2. Across the six included studies (n = 68), all outcomes (ΔrCBV, reader-rated margin delineation, CNR/SNR, and safety) were graded as low certainty. For the ΔrCBV (two studies, n = 33; Dósa [28], Stoller [29]), the consistency of direction favored NP-CAs, but imprecision was severe due to sample size and wide variance. Margin delineation (five studies, n = 42) showed uniform qualitative improvement, with USPIOs persisting up to 36 h; however, it was downgraded for imprecision, as most datasets involved ≤ 9 patients and lacked quantitative precision metrics. CNR/SNR gains (three studies, n ≈ 35) were downgraded for inconsistency (heterogeneous metrics and acquisition settings) and imprecision. Safety reporting (in all six studies) identified no serious adverse events but possibly under-reported, and small aggregates prompted downgrading for imprecision and suspected reporting bias. No outcome reached high or moderate certainty, primarily because of small studies, non-consecutive recruitment, heterogeneity in agents and MRI protocols, and incomplete adverse-event documentation. These certainty ratings contextualize the directionally consistent but methodologically constrained signals observed in this review.

**Table 2.** Certainty of evidence (GRADE)

| Outcome | No. of studies (n) | Study designs | Risk of Bias | Inconsistency | Indirectness | Imprecision | Overall GRADE certainty | Summary of findings (NP-CA vs Gd) |
| --- | --- | --- | --- | --- | --- | --- | --- | --- |
| ΔrCBV (relative cerebral blood volume) | 2 studies (Dósa 2011; Stoller 2023), n=33 | Cohort | Not serious (low to unclear RoB) | Not serious (direction consistent) | Not serious (direct comparison with Gd) | Serious (small total n; wide CI) | Low | Ferumoxytol yielded ~18–25% higher rCBV than Gd in DSC; measurements stable despite leakage. |
| Margin delineation (reader-rated/visual) | 5 studies (Enochs; Varallyay; Taschner; Hunt; Stoller), n=42 | Case series; cohort | Not serious (low RoB) | Not serious (clearer/thinner rims vs Gd across studies) | Not serious | Serious (most samples 2–9; limited quantitative precision measures) | Low | USPIO rims sharper and persist to ~36 h; often extend beyond Gd enhancement; histology shows immune-rich margins. |
| CNR/SNR improvements | 3 studies, n~35 | Case series | Not serious | Serious (different metrics; heterogeneous effect sizes) | Not serious | Serious (small n) | Low | Directionally higher CNR/SNR with NP-CAs, but magnitude uncertain due to varied acquisition and definitions. |
| Safety (adverse events) | 6 studies, n=68 | Case report; case series; cohort | Not serious (mostly low RoB; some incomplete AE reporting) | Not serious | Not serious | Serious (few participants; likely underreporting) | Low | No serious adverse events reported; infusion reactions rarely detailed; no signal for harm in limited data. |
| Abbreviations: NP-CA = nanoparticle contrast agent; Gd = gadolinium; USPIO = ultrasmall superparamagnetic iron oxide; DSC = dynamic susceptibility contrast; rCBV = relative cerebral blood volume; CNR = contrast-to-noise ratio; SNR = signal-to-noise ratio; AE = adverse event; RoB = risk of bias. |  |  |  |  |  |  |  |  |

## Discussion

### Principal Findings

Meta-analytic synthesis of fixed-dose regimens across all six eligible studies (n = 68) demonstrated two distinct dosing clusters (one near 2.6 mg Fe/kg (commonly associated with ferumoxtran-10 protocols) and another at 7.0 mg Fe/kg (used predominantly with ferumoxytol). The common-effect pooled mean dose was 4.65 mg Fe/kg (95% CI 4.62–4.67), whereas the random-effects model (REML + Hartung–Knapp adjustment) yielded 3.82 mg Fe/kg (95% CI 1.16–6.48). Between-study heterogeneity was extreme (I² = 100%, τ² = 6.4166), reflecting not random variation but protocol-driven bimodality. This pronounced dose difference suggests that regimen selection is a major determinant of the imaging phenotype, potentially influencing susceptibility rim conspicuity, DSC perfusion stability, and the balance between vascular versus cellular/matrix targeting. These findings highlight the need for controlled dose response trials before widespread protocol standardization.

In this systematic review, iron-oxide nanoparticle contrast agents (NP-CAs), primarily ferumoxtran-10 and ferumoxytol, consistently enhanced neuro-oncologic MRI performance [27–32]. Across heterogeneous platforms (0.15–3 T) and designs, two principal benefits emerged:

1. Sharper, persistent susceptibility rims on T2/T2*/GRE/SWI sequences often extend beyond T1-Gd enhancement and last up to ∼ 36 h post-injection [27,28,30–32].
2. Compared with that of gadolinium, the ΔrCBV of DSC perfusion is greater and more stable, with fewer leakage artifacts [28,29,36].

A critical methodological finding of this review is that none of the prespecified imaging outcomes reached the minimum threshold for meta-analysis. ΔrCBV values were available from a single study (Dósa 2011), CNR/SNR either lacked variance or were reported in incomparable units, and all reader-rated margin measures were qualitative. This left per-protocol dose as the only analyzable parameter. Because each study administered a uniform fixed dose to all participants, within-study variability reflects protocol noise rather than biological variance, permitting a justified single-arm dose meta-analysis. The resulting bimodal distribution (2.6 vs. 7.0 mg/kg) reflects true protocol heterogeneity rather than statistical artifact, providing a mechanistic explanation for divergent imaging phenotypes across studies.

A key methodological finding of this review is that none of the prespecified imaging outcomes met the minimum statistical threshold for quantitative synthesis. Extractable ΔrCBV values with variance were available from only a single study (Dósa 2011); CNR and SNR datasets were either absent or reported without variance; and all reader-rated margin assessments were qualitative across studies. As a result, no imaging outcome achieved the ≥3 comparable datasets required under PRISMA 2020 and the PROSPERO-registered protocol. This left the per-protocol nanoparticle iron dose as the only analyzable parameter. Because dosing was fixed within each study, within-study variability reflects protocol noise rather than biological variation, enabling a justified single-arm meta-analysis. The distinctly bimodal pooled dose distribution (≈2.6 mg/kg vs. ≈7.0 mg/kg) reflects true protocol heterogeneity and plausibly explains the divergent susceptibility- and perfusion-based imaging phenotypes observed across studies.

### Agreement and Divergence with Previous Literature

Our results align with prior clinical series [27,28,30–32] reporting delayed USPIO rims that precisely delineate infiltrative tumor borders and, in some cases, highlight lesion extent beyond Gd enhancement. Stoller [29] linked these rims to immune-cell-rich microenvironments via stereotactic biopsy, supporting a biological basis (macrophage/inflammation) rather than purely vascular leakage.

### Key divergences include the following

- T1 crispness: Dósa [28] reported sharper T1 margins with Gd, even though ferumoxytol provided superior DSC perfusion metrics.
- Lesion detectability: Enochs [32] described Gd-positive/USPIO-negative lesions, possibly reflecting minimal macrophage infiltration.
- Agent chemistry: Varallyay [31] reported negligible uptake of ferumoxides compared with ferumoxtran-10, underscoring the relevance of particle size, coating, and surface chemistry.

### Mechanistic considerations

Early-phase ferumoxytol remains intravascular, stabilizing DSC-derived rCBV values and reducing the leakage sensitivity inherent to Gd [28,29,36]. Ferumoxtran-10, in contrast, progressively localizes to interstitial and intracellular compartments, producing conspicuous susceptibility rims at 24 h [27,30,31]. Higher field strengths (3 T) increase the rim contrast-to-edema ratios [29]. The strong bimodality in pooled dose data supports a hypothesized dose response effect that remains untested in controlled trials.

### Protocol Implications

Three actionable parameters emerge from the synthesis:

1. Sequence pairing: Combine susceptibility-sensitive GRE/SWI with T1-SE/TSE for simultaneous rim/core assessment [27,28,30,31].
2. Timing: Acquire early images for perfusion; perform susceptibility imaging at 24 h to maximize rim conspicuity while avoiding blooming/decaying from very late acquisitions [27–32].
3. Dosing: Harmonized weight-based dosing (target range 2–5 mg Fe/kg) is adopted to minimize body-habitus--related variability, and protocols-driven extremes (2.6 vs. 7 mg/kg) are avoided unless justified by investigational aims [Dosage meta-analysis, this review]. Slow infusion rates may mitigate infusion reactions [28].

### Clinical Applications

#### The evidence converges on three potential indications

- Mapping tumor infiltration: USPIO rims extending into FLAIR-abnormal zones may define macrophage-rich invasive margins [27,29].
- Progression vs. radiation necrosis: Absent USPIO uptake in presumed necrosis [27] and ferumoxytol identification of inflammatory signatures [28,29] can clarify ambiguous post-treatment imaging.
- Perfusion quantification in leaky tumors: Ferumoxytol offers stable, high rCBV values when Gd-DSC is compromised by BBB disruption [28,36].

These features could support broader resection margins, refine radiotherapy contours, and reduce diagnostic uncertainty—pending protocol standardization.

### Risk of bias and certainty

Using the JBI tool [25,26], five studies were low risk [27,28,30–32], whereas Stoller [29] reported “unclear” risk due to incomplete reporting and baseline imbalances. Common limitations:

- small sample sizes (n=2–26 per study; total n=68),
- non-consecutive enrollment,
- incomplete demographic/follow-up data,
- inconsistent adverse-event definitions.

According to our GRADE assessment, certainty across all primary and secondary outcomes remains low, primarily owing to imprecision, heterogeneity, and design limitations.

### Safety and Operational Context

No serious adverse events were reported [27–32]; however, safety surveillance was inconsistent, and mild infusion reactions could be under-reported [28]. Operational deployment is constrained by off-label regulatory status, requiring explicit informed consent, pharmacy coordination, and quality-management protocols.

### Limitations

- Quantitative synthesis was possible only for pooled dosing, not for primary imaging outcomes, owing to incomplete variance reporting and heterogeneity in readouts (ΔrCBV, CNR, qualitative margins).
- The bimodal dose distribution compromises the extrapolation of performance metrics.
- Imaging field strengths, sequence types, and timing vary widely.
- Only one study [29] provided histopathology-linked imaging validation.
- Publication bias cannot be excluded, as all studies reported favorable findings.
- Additionally, selective reporting of positive findings in the literature suggests a risk of publication bias, which may overestimate the magnitude of observed benefits.

### Future Directions

1. Trial Design: Multicenter RCTs or well-designed within-patient crossover studies directly comparing NP-CAs with Gd under matched MRI conditions [23,24].
2. Protocol Harmonization: Address bimodal dosing by consensus on weight-adjusted regimens (2–5 mg Fe/kg), standardized infusion rates, and fixed imaging windows.
3. Outcome Reporting – Require variance-reported quantitative metrics and standardized AE frameworks; commit to publishing negative results.
4. Biopsy-Anchored Validation: Systematically sample rim/core regions to confirm cellular/molecular correlates across tumor subtypes [29].
5. Clinical Integration: Evaluate workflow impact in surgical navigation and RT planning; maintain longitudinal safety registries.
6. Advanced Analytics: Develop AI/radiomics models using harmonized acquisition protocols [3,6,7,11,12] to derive generalizable predictive biomarkers. Standardized imaging datasets from future trials will also be essential for training and validating AI-and radiomics-based tools for automated tumor margin assessment.

### Conclusion

Iron-oxide NP-CAs add biologically relevant contrast beyond Gd, offering both stable perfusion metrics and delayed susceptibility rims mapping macrophage-rich infiltration. Pooled dose analysis confirmed heterogeneity driven by protocol-level decisions—a modifiable factor. Rigorous, standardized, biopsy-validated trials are essential to translate these promising findings into practice-changing evidence.

## Data Availability

The data that support the findings of this study are available from the corresponding author upon reasonable request.

## Declarations

### Human Ethics

This is a systematic review and meta-analysis study, and it is deemed exempt from obtaining ethical approval.

## Acknowledgement

We thank colleagues at the Shohada Tajrish Comprehensive Neurosurgical Center of Excellence (Shahid Beheshti University of Medical Sciences), the School of Medicine (Tehran University of Medical Sciences), and our collaborating Student Research Committees (Hormozgan, Semnan, and Larestan Universities of Medical Sciences) for administrative and logistical support.

## AI Use Disclosure

Artificial intelligence tool(Chat GPT 5.1) were used for refinement and editing of language in this manuscript. These tools assisted with grammar checking, phrasing suggestions, and improvements to clarity and readability. No AI tools were used to generate any scientific content, conduct data analysis, or create new text from prompts. All conceptualization, data interpretation, and writing of original content were performed entirely by the authors.

