## supplementary file 5 for "Iron-Oxide Nanoparticle MRI for Human Brain Tumors: A Systematic Review and Protocol-Level Meta-analysis of Administered Doses Across Ferumoxtran-10, Ferumoxytol, and Ferumoxides"

|  | Taschner et al.et al | | | Varallyay et al. | | | Enochs et al. | | | Dósa et al. | | |
| --- | --- | --- | --- | --- | --- | --- | --- | --- | --- | --- | --- | --- |
|  | R1=NSD | R2=AK | R3=ARv | R1=NSD | R2=AK | R3=ARv | R1=NSD | R2=AK | R3=ARv | R1=NSD  R2=AK |  | R3=ARv |
| Were there clear criteria for inclusion in the case series? | + | + | + | + | + | + | + | + | + | + | + | + |
| Was the condition measured in a standard, reliable way for all participants included in the case series? | + | + | + | + | + | + | + | + | + | + | + | + |
| Were valid methods used for identification of the condition for all participants included in the case series? | + | + | + | + | + | + | + | + | + | + | + | + |
| Did the case series have consecutive inclusion of participants? | + | + | + | × | + | - | × | + | - | × | + | N/A |
| Did the case series have complete inclusion of participants? | + | + | + | + | + | + | + | - | - | - | + | - |
| Was there clear reporting of the demographics of the participants in the study? | + | + | + | + | + | + | + | + | + | + | + | + |
| Was there clear reporting of clinical information of the participants? | + | + | + | + | + | + | + | - | + | + | + | + |
| Were the outcomes or follow up results of cases clearly reported? | + | + | + | + | + | + | + | + | + | + | + | + |
| Was there clear reporting of the presenting site(s)/clinic(s) demographic information? | + | - | + | + | - | + | + | - | - | + | + | + |
| Was statistical analysis appropriate? | + | + | + | + | + | + | + | + | + | + | + | + |

1. **JBI checklist for case series**

Yes +

No -

Unclear ×

Not Applicable N/A

2. **JBI checklist for case series**

|  | Stoller et al. | | |
| --- | --- | --- | --- |
|  | R2=AK  R1=NSD |  | R3=ARv |
| Were the two groups similar and recruited from the same population? | × | + | - |
| Were the exposures measured similarly to assign people to both exposed and unexposed groups? | × | + | + |
| Was the exposure measured in a valid and reliable way? | + | + | + |
| Were confounding factors identified? | × | + | + |
| Were strategies to deal with confounding factors stated? | - | + | + |
| Were the groups/participants free of the outcome at the start of the study (or at the moment of exposure)? | × | + | + |
| Were the outcomes measured in a valid and reliable way? | + | + | + |
| Was the follow-up time reported and sufficient to be long enough for outcomes to occur? | N/A | × | × |
| Was follow-up complete, and if not, were the reasons for loss to follow-up described and explored? | N/A | × | × |
| Were strategies to address incomplete follow-up utilized? | N/A | - | - |
| Was appropriate statistical analysis used? | + | + | + |

3. **JBI checklist for cohorts**

|  | Hunt MA, Bago AG, Neuwelt EA | | |
| --- | --- | --- | --- |
|  | R1=NSD | R2=AK | R3=ARv |
| Were patient’s demographic characteristics clearly described? | + | + | + |
| Was the patient’s history clearly described and presented as a timeline? | + | + | + |
| Was the current clinical condition of the patient on presentation clearly described? | × | + | + |
| Were diagnostic tests or assessment methods and the results clearly described? | + | + | + |
| Was the intervention(s) or treatment procedure(s) clearly described? | + | + | + |
| Was the post-intervention clinical condition clearly described? | × | + | × |
| Were adverse events (harms) or unanticipated events identified and described? | - | - | - |
| Does the case report provide takeaway lessons? | + | + | + |
