## supplementary file 2 for "Iron-Oxide Nanoparticle MRI for Human Brain Tumors: A Systematic Review and Protocol-Level Meta-analysis of Administered Doses Across Ferumoxtran-10, Ferumoxytol, and Ferumoxides"

Level Meta-analysis of Administered Doses Across Ferumoxtran10, Ferumoxytol, and Ferumoxides. PROSPERO 202
CRD420251106029. Available from https://www.crd.york.ac.uk/PROSPERO/view/CRD420251106029.

REVIEW

### REVIEW TITLE AND BASIC DETAILS

#### Review title

Ironoxide Nanoparticle MRI for Human Brain Tumors: A Systematic Review and Protocol-Level Meta-analysis of Adminis
Across Ferumoxtran10, Ferumoxytol, and Ferumoxides

#### Condition or domain being studied

*Brain neoplasm; Mri Of Brain With T2 Mapping*

#### Rationale for the review

Conventional gadolinium-based contrast agents (GBCAs) have limited ability to depict infiltrative tumour borders becaus
blood–brain-barrier leakage. Nanoparticle-based agents (e.g., super-paramagnetic iron-oxide, manganese, gold, hafniun
formulations) exhibit prolonged circulation, active or size-dependent tumour uptake and higher r* relaxivity, potentially pro
margin definition. Individual pre-clinical and early-phase clinical studies report heterogeneous results; a rigorous synthes
quantify their true added value and identify moderators (NP type, field strength, tumour histology, etc.).

#### Review objectives

1. Quantify the improvement in margin-delineation accuracy (e.g., Dice similarity coefficient, Hausdorff distance) obtaine
nanoparticle-based contrast vs (a) standard GBCAs, (b) non-contrast MRI, or (c) histopathology.

2. Compare contrast-to-noise ratio (CNR) and signal-to-noise ratio (SNR) between nanoparticle-enhanced and conventio

3. Explore how agent composition, surface functionalisation, dose, MRI field strength and study design influence the effe

4. Summarise safety/adverse-event data where reported.

#### Keywords

Brain tumour; Nanoparticles; Contrast agents; MRI; margin delineation

#### Country

Iran (Islamic Republic of)

### ELIGIBILITY CRITERIA

#### Population

*Included*

• Human patients with any primary or secondary brain tumour imaged by MRI.

• No age, sex, or tumour-grade limits.

*Excluded*

• In-vitro phantom studies and animal studies.

• Imaging of non-brain tumours.

#### Intervention(s) or exposure(s)

*Included*

*Mri Of Brain With T2 Mapping; Magnetic Resonance Imaging With Contrast*

Administration of any nanoparticle-based MRI contrast agent (iron-oxide, manganese, gold, silica, carbon, polymeric, lipi
targeted NPs).

*Excluded*

Non-nanoparticulate contrast agents (e.g., gadolinium chelates), unless used solely as comparator.

#### Comparator(s) or control(s)

*Included*

Pre-clinical controlled or uncontrolled animal studies, diagnostic accuracy or effectiveness studies (RCTs, cohort, case–c
sectional) in humans.

*Excluded*
Case reports (less than 5 subjects), reviews, editorials, abstracts without full data.

#### Context
Any imaging centre or laboratory worldwide; in vivo MRI at any field strength

### SIMILAR REVIEWS

#### Check for similar records already in PROSPERO
*PROSPERO identified a number of existing PROSPERO records that were similar to this one (last check made on 16 Ju are shown below along with the reasons given by that the review team for the reviews being different and/or proceeding.*

- Gadolinium-based contrast agents and t1-hypersignals in human brain: a systematic review [published 17 April 2016 [CRD42016037902]. The review was judged **not to be similar**
- Efficacy and safety of half-dose Gadopiclenol versus full dose Gadobutrol for Contrast-Enhanced MRI of the body a nervous system: A Meta-Analysis [published 27 December 2023] [CRD42023494255]. The review was judged **not t**
- Efficacy and safety of contrast media used in cardiac magnetic resonance imaging (MRI): a systematic review [publi 2024] [CRD42024539482]. The review was judged **not to be similar**

### TIMELINE OF THE REVIEW

#### Date of first submission to PROSPERO
16 July 2025

#### Review timeline
Start date: 16 July 2025. End date: 16 October 2025.

#### Date of registration in PROSPERO
17 July 2025

### AVAILABILITY OF FULL PROTOCOL

#### Availability of full protocol
A full protocol has not been written.

### SEARCHING AND SCREENING

#### Search for unpublished studies
Only published studies will be sought.

#### Main bibliographic databases that will be searched
The main databases to be searched are *MEDLINE*, *PubMed*, *SCI - Science Citation Index* and *Scopus*.

#### Search language restrictions
There are no language restrictions.

#### Search date restrictions
There are no search date restrictions.

#### Other methods of identifying studies
Other studies will be identified by: *looking through all the articles that cite the papers included in the review ("snowballing list checking*.

#### Link to search strategy
A full search strategy has been uploaded to PROSPERO. The PDF may be accessed through this link https://www.crd.york.ac.uk/PROSPEROFILES/bdf966d9c21bf8b58f612bb6883c35d2.pdf.

#### Study risk of bias or quality assessment

Risk of bias will be assessed using:

JBI Critical Appraisal Tools

Data will be assessed independently by at least two people (or person/machine combination) with a process to resolve d

Additional information will be sought from study investigators if required information is unclear or unavailable in the study publications/reports.

#### Reporting bias assessment

Risk of bias due to missing results will be assessed

#### Certainty assessment

Certainty of findings will not be assessed

### OUTCOMES TO BE ANALYSED

#### Main outcomes

primary:

• Dice similarity coefficient (tumour vs ground truth)

• Hausdorff distance / Boundary distance error

secondary:

• Contrast-to-noise ratio (lesion vs normal parenchyma)

• Signal-to-noise ratio

• Extent-of-resection accuracy (for surgical studies)

• Adverse events (nephrotoxicity, inflammatory response).

#### Additional outcomes

There are no additional outcomes.

### PLANNED DATA SYNTHESIS

#### Strategy for data synthesis

No formal data synthesis is planned - data will be described but not combined.

### CURRENT REVIEW STAGE

#### Stage of the review at this submission

| Review stage | Started | Completed |
|---|---|---|
| Pilot work | ✓ | |
| Formal searching/study identification | ✓ | |
| Screening search results against inclusion criteria | | |
| Data extraction or receipt of IPD | | |
| Risk of bias/quality assessment | | |
| Data synthesis | | |

#### Review status

The review is currently planned or ongoing.

#### Publication of review results

Results of the review will be published in English.

#### Review affiliation

Functional Neurosurgery Research Center, Shohada Tajrish Comprehensive Neurosurgical Center of Excellence, Shahid
University of Medical Sciences, Tehran, Iran

#### Funding source

Review has no specific/external funding but is supported by guarantor/review team (non-commercial) institutions.

#### Peer review

There has been no peer review of this planned review.

### ADDITIONAL INFORMATION

#### Review conflict of interest

Declared individual interests are recorded under team member details.. No additional interests are recorded for this revie

#### Medical Subject Headings

Brain Neoplasms; Contrast Media; Humans; Magnetic Resonance Imaging; Nanoparticles

#### Revision note

No preview available

#### PROSPERO version history

- Version 1.0, published 17 Jul 2025

#### Disclaimer

The content of this record displays the information provided by the review team. PROSPERO does not peer review regis
endorse their content.

PROSPERO accepts and posts the information provided in good faith; responsibility for record content rests with the rev
guarantor for this record has affirmed that the information provided is truthful and that they understand that deliberate pro
inaccurate information may be construed as scientific misconduct.

PROSPERO does not accept any liability for the content provided in this record or for its use. Readers use the informatio
record at their own risk.

Any enquiries about the record should be referred to the named review contact
