## supplementary file 3 for "Iron-Oxide Nanoparticle MRI for Human Brain Tumors: A Systematic Review and Protocol-Level Meta-analysis of Administered Doses Across Ferumoxtran-10, Ferumoxytol, and Ferumoxides"

| 12 July 2025 | | |
| --- | --- | --- |
| **Pubmed** | ( ("margin*" OR "border*" OR "edge*" OR "delineat*" OR "rim" OR "boundary*" OR "outline*" OR "contour*" OR "periphery" OR "margin delineation" OR "tumor margin" OR "tumor border") AND ("contrast" OR "contrast agent*" OR "contrast enhancement" OR "contrast media" OR "contrast material" OR "contrast dye" OR "contrast medium" OR "paramagnetic agent*" OR "imaging agent*") AND (nanoparticle* OR "nano particle*" OR nanotech* OR "nanotechnology" OR "nanostructure*" OR "quantum dot*" OR "superparamagnetic iron oxide" OR SPIO OR "ultrasmall superparamagnetic iron oxide" OR USPIO OR "iron oxide nanoparticle*" OR "magnetic nanoparticle*" OR "magnetite nanoparticle*" OR "liposomal nanoparticle*" OR "polymeric nanoparticle*" OR "gold nanoparticle*" OR "metallic nanoparticle*" OR "carbon nanoparticle*" OR "nanocrystal*" OR "nanocomposite*") AND (MRI OR "magnetic resonance imaging" OR "magnetic resonance" OR "MR imaging" OR "magnetic resonance image*" OR "magnetic resonance scan*" OR "MR scan*") AND ("brain tumor*" OR glioma* OR "glioblastoma*" OR "astrocytoma*" OR "oligodendroglioma*" OR "ependymoma*" OR "medulloblastoma*" OR "brain neoplasm*" OR "central nervous system tumor*" OR "CNS tumor*" OR "meningioma*" OR "primary brain tumor*" OR "intracranial tumor*") ) | 70 |
| **Scopus** | ( TITLE-ABS-KEY ( ( "margin*" OR "border*" OR "edge*" OR "delineat*" OR "rim" OR "boundary*" OR "outline*" OR "contour*" OR "periphery" OR "tumor margin" OR "tumor border" ) AND ( "contrast" OR "contrast agent*" OR "contrast enhancement" OR "contrast media" OR "contrast material" OR "contrast dye" OR "paramagnetic agent*" OR "imaging agent*" ) AND ( nanoparticle* OR "nano particle*" OR nanotech* OR "nanotechnology" OR "nanostructure*" OR "quantum dot*" OR "superparamagnetic iron oxide" OR SPIO OR USPIO OR "iron oxide nanoparticle*" OR "magnetic nanoparticle*" OR "magnetite nanoparticle*" OR "liposomal nanoparticle*" OR "polymeric nanoparticle*" OR "gold nanoparticle*" OR "metallic nanoparticle*" OR "carbon nanoparticle*" OR "nanocrystal*" OR "nanocomposite*" ) AND ( MRI OR "magnetic resonance imaging" OR "magnetic resonance" OR "MR imaging" OR "magnetic resonance image*" OR "magnetic resonance scan*" OR "MR scan*" ) AND ( "brain tumor*" OR glioma* OR "glioblastoma*" OR "astrocytoma*" OR "oligodendroglioma*" OR "ependymoma*" OR "medulloblastoma*" OR "brain neoplasm*" OR "central nervous system tumor*" OR "CNS tumor*" OR "meningioma*" OR "primary brain tumor*" OR "intracranial tumor*" ) ) ) | 102 |
| **WOS** | TS=(  ("margin*" OR "border*" OR "edge*" OR "delineat*" OR "rim" OR "boundary*" OR "outline*" OR "contour*" OR "periphery" OR "tumor margin" OR "tumor border")  AND  ("contrast" OR "contrast agent*" OR "contrast enhancement" OR "contrast media" OR "contrast material" OR "contrast dye" OR "paramagnetic agent*" OR "imaging agent*")  AND  (nanoparticle* OR "nano particle*" OR nanotech* OR "nanotechnology" OR "nanostructure*" OR "quantum dot*" OR "superparamagnetic iron oxide" OR SPIO OR USPIO OR "iron oxide nanoparticle*" OR "magnetic nanoparticle*" OR "magnetite nanoparticle*" OR "liposomal nanoparticle*" OR "polymeric nanoparticle*" OR "gold nanoparticle*" OR "metallic nanoparticle*" OR "carbon nanoparticle*" OR "nanocrystal*" OR "nanocomposite*")  AND  (MRI OR "magnetic resonance imaging" OR "magnetic resonance" OR "MR imaging" OR "magnetic resonance image*" OR "magnetic resonance scan*" OR "MR scan*")  AND  ("brain tumor*" OR glioma* OR "glioblastoma*" OR "astrocytoma*" OR "oligodendroglioma*" OR "ependymoma*" OR "medulloblastoma*" OR "brain neoplasm*" OR "central nervous system tumor*" OR "CNS tumor*" OR "meningioma*" OR "primary brain tumor*" OR "intracranial tumor*")  ) | 83 |
| **Cochrane** | (margin* OR border* OR edge* OR delineat* OR rim OR boundary* OR outline* OR contour* OR periphery OR "margin delineation" OR "tumor margin" OR "tumor border") AND (contrast OR "contrast agent*" OR "contrast enhancement" OR "contrast media" OR "contrast material" OR "contrast dye" OR "contrast medium" OR "paramagnetic agent*" OR "imaging agent*")AND (nanoparticle* OR "nano particle*" OR nanotech* OR nanotechnology OR nanostructure* OR "quantum dot*" OR "superparamagnetic iron oxide" OR spio OR "ultrasmall superparamagnetic iron oxide" OR uspio OR "iron oxide nanoparticle*" OR "magnetic nanoparticle*" OR "magnetite nanoparticle*" OR "liposomal nanoparticle*" OR "polymeric nanoparticle*" OR "gold nanoparticle*" OR "metallic nanoparticle*" OR "carbon nanoparticle*" OR nanocrystal* OR nanocomposite*)AND (MRI OR "magnetic resonance imaging" OR "magnetic resonance" OR "MR imaging" OR "magnetic resonance image*" OR "magnetic resonance scan*" OR "MR scan*") AND ("brain tumor*" OR glioma* OR "glioblastoma*" OR astrocytoma* OR oligodendroglioma* OR ependymoma* OR medulloblastoma* OR "brain neoplasm*" OR "central nervous system tumor*" OR "CNS tumor*" OR meningioma* OR "primary brain tumor*" OR "intracranial tumor*") in Title Abstract Keyword | 2 |
| **Embase** | (margin*:ti,ab,kw OR border*:ti,ab,kw OR edge*:ti,ab,kw OR delineat*:ti,ab,kw OR rim:ti,ab,kw OR boundary*:ti,ab,kw OR outline*:ti,ab,kw OR contour*:ti,ab,kw OR periphery:ti,ab,kw OR 'margin delineation':ti,ab,kw OR 'tumor margin':ti,ab,kw OR 'tumor border':ti,ab,kw) AND (contrast:ti,ab,kw OR 'contrast agent*':ti,ab,kw OR 'contrast enhancement':ti,ab,kw OR 'contrast media':ti,ab,kw OR 'contrast material':ti,ab,kw OR 'contrast dye':ti,ab,kw OR 'contrast medium':ti,ab,kw OR 'paramagnetic agent*':ti,ab,kw OR 'imaging agent*':ti,ab,kw) AND (nanoparticle*:ti,ab,kw OR 'nano particle*':ti,ab,kw OR nanotech*:ti,ab,kw OR nanotechnology:ti,ab,kw OR nanostructure*:ti,ab,kw OR 'quantum dot*':ti,ab,kw OR 'superparamagnetic iron oxide':ti,ab,kw OR spio:ti,ab,kw OR 'ultrasmall superparamagnetic iron oxide':ti,ab,kw OR uspio:ti,ab,kw OR 'iron oxide nanoparticle*':ti,ab,kw OR 'magnetic nanoparticle*':ti,ab,kw OR 'magnetite nanoparticle*':ti,ab,kw OR 'liposomal nanoparticle*':ti,ab,kw OR 'polymeric nanoparticle*':ti,ab,kw OR 'gold nanoparticle*':ti,ab,kw OR 'metallic nanoparticle*':ti,ab,kw OR 'carbon nanoparticle*':ti,ab,kw OR nanocrystal*:ti,ab,kw OR nanocomposite*:ti,ab,kw) AND (mri:ti,ab,kw OR 'magnetic resonance imaging':ti,ab,kw OR 'magnetic resonance':ti,ab,kw OR 'mr imaging':ti,ab,kw OR 'magnetic resonance image*':ti,ab,kw OR 'magnetic resonance scan*':ti,ab,kw OR 'mr scan*':ti,ab,kw) AND ('brain tumor*':ti,ab,kw OR glioma*:ti,ab,kw OR 'glioblastoma*':ti,ab,kw OR astrocytoma*:ti,ab,kw OR oligodendroglioma*:ti,ab,kw OR ependymoma*:ti,ab,kw OR medulloblastoma*:ti,ab,kw OR 'brain neoplasm*':ti,ab,kw OR 'central nervous system tumor*':ti,ab,kw OR 'cns tumor*':ti,ab,kw OR meningioma*:ti,ab,kw OR 'primary brain tumor*':ti,ab,kw OR 'intracranial tumor*':ti,ab,kw) | 67 |
| **Duplicate** | | 149 |
| **Final screening** | | 175 |
